# Characteristics and Outcomes of Gene-Elusive Dilated Cardiomyopathy

**DOI:** 10.64898/2026.06.17.26355852

**Authors:** Douglas E Cannie, Athanasios Bakalakos, Petros Syrris, Alexandros Protonotarios, Massimiliano Lorenzini, Oliver Guttmann, Constantinos O’Mahony, Konstantinos Savvatis, Neha Sekhri, Saidi A Mohiddin, Juan Pablo Kaski, Luis R Lopes, Perry M Elliott

## Abstract

**Background and Aims:** Genetic testing in dilated cardiomyopathy (DCM) guides risk stratification and family screening. Likely pathogenic or pathogenic (LP/P) variants are identified in approximately one-third of patients, leaving many without a genetic diagnosis. Cohort studies suggest that ‘gene-elusive’ patients have a lower risk of adverse events. This study aims to better characterise this group and identify factors associated with adverse outcomes.

**Methods:** Consecutive and unrelated DCM patients undergoing genetic testing and returning no LP/P variants were retrospectively recruited and compared to two control cohorts of DCM patients carrying LP/P variants in *LMNA* and *TTN* for a primary composite endpoint of end-stage heart failure (ESHF) or malignant ventricular arrhythmia (MVA).

**Results:** Among patients without prior MVA, the composite endpoint occurred in 36/423 (8.5%) gene-elusive, 14/39 (35.9%) *LMNA* and 11/100 (11%) *TTN* cardiomyopathy patients (log-rank p < 0.001 for *LMNA* vs gene-elusive and *LMNA* vs *TTN*; p = 0.96 for *TTN* vs gene-elusive).

For gene-elusive patients, lower left ventricular ejection fraction, larger left ventricular internal diameter in diastole, absence of LBBB and ventricular ectopy on ECG were independent predictors of the primary endpoint. Gene-elusive patients with LBBB had less atrial arrhythmia and a lower burden of ventricular ectopy at baseline and a low risk of the primary composite endpoint (HR 0.3 [0.1-0.8], p-value 0.01).

**Conclusions:** Gene-elusive DCM patients have a risk of adverse events similar to *TTN* cardiomyopathy. Gene-elusive patients with LBBB form a particularly low-risk subgroup, likely reflecting a distinct aetiology of left ventricular systolic dysfunction.

## Introduction

Dilated cardiomyopathy (DCM) is a disease of heart muscle characterised by left ventricular systolic dysfunction (LVSD) and dilatation unexplained by coronary artery disease or loading conditions. Recent decades have seen significant progress in understanding the genetic factors underpinning DCM with data showing that 19-37% of patients have disease caused by rare pathogenic variants in single genes. Increasingly, evidence demonstrates that specific genotypes cause distinct phenotypes with different risk profiles that can be used to guide therapy and plan long-term clinical surveillance.^1–3^

In contrast, some studies suggest that gene-elusive status confers a much lower risk of progression to heart failure and malignant ventricular arrhythmia than that observed in patients with Mendelian forms of DCM; however, this has not been systematically evaluated. If this hypothesis is correct, a more conservative approach to disease surveillance might be appropriate in gene-elusive DCM patients.

This single-centre cohort study aimed to characterise the risk profile of patients with gene-elusive DCM compared with those harbouring *LMNA* or *TTN* variants, and to identify baseline clinical features associated with adverse outcomes.

## Methods

### Study Design and Cohort Composition

This was a single centre, retrospective longitudinal cohort study. The study conforms to the principles of the Declaration of Helsinki^4^ and participants provided written informed consent for genetic testing. Local ethical approval for anonymised patient data collection and analysis was obtained (19/WS/0100). Patient level data will not be made available as consent was not sought for public dissemination and due to concerns that information could be used to identify individuals.

Consecutive patients seen at Barts Heart Centre, London with DCM (defined as LVEF < 50% in the absence of coronary artery disease or abnormal loading conditions sufficient to cause the phenotype), that had genetic testing between 2015 and 2019 and did not return a likely pathogenic or pathogenic (LP/P) variant (‘gene-elusive’) were recruited from the University College London (UCL) and Barts Heart Centre cardiomyopathy database.

Next-generation targeted sequencing using multigene panels of at least 81 genes (Supplementary Table 1) was performed at Health in Code (HiC, https://healthincode.com/en/), an accredited diagnostic laboratory with results interpreted according to modified American College of Medical Genetics (ACMG) criteria.^5^ A proband was defined as an index patient with DCM. No gene-elusive patients in this study were from the same family.

Variants were initially classified by HiC using modified American College of Medical Genetics (ACMG) criteria. For the purposes of this study, all variants were reassessed in November and December 2025 using ACMG criteria.^5^ Where additional evidence of pathogenicity (e.g. family cosegregation) was available, variants were reclassified accordingly.

For comparator cohorts, the UCL and Barts Heart Centre database was reviewed for all carriers of LP/P variants in *TTN* and *LMNA*. To define clinically affected patients, we applied a penetrance definition designed to capture clinically actionable disease features. Specifically, disease penetrance was defined by the presence of one or more of the following: (1) LVEF < 50% on TTE or cardiac MRI; (2) non-sustained ventricular tachycardia (NSVT; defined as 3 or more consecutive ventricular beats at ≥120bpm) or premature ventricular complex (PVC) count > 500/24hours on Holter monitoring^6–8^, (3) non-ischaemic late gadolinium enhancement (LGE) on cardiac MRI, (4) early-onset atrial fibrillation or flutter (< 40 years of age), and (5) sino-atrial node disease or high-degree AV block requiring a permanent pacemaker (PPM). To ensure consistency across cohorts, the penetrance definition used for *TTN* and *LMNA* variant-carriers was applied to gene-elusive DCM patients in order to determine those clinically affected at baseline.

*TTN* and *LMNA* variant pathogenicity was manually reviewed in December 2025 using ACMG criteria^5^ in conjunction with Gnomad (v4.1.0)^9^ and ClinVar.^10^ Where additional evidence of pathogenicity (e.g. familial cosegregation) was available, variants were reclassified accordingly (Supplementary Table 2).

### Data Collection and Study Variables

Study data were collected and managed using a REDCap (Research Electronic Data Capture), database hosted at UCL.^11^ Baseline demographics, family history, comorbidities, symptoms, 12-lead electrocardiogram (ECG), transthoracic echocardiogram (TTE), ambulatory ECG recordings and cardiac magnetic resonance (CMR) scan data were collected from clinical records. The baseline phenotypic data comprised the primary dataset used for outcomes analyses.

Left bundle branch block (LBBB) was defined as per AHA/ACCF/HRS recommendations to include a QRS duration of greater than or equal to 120ms in addition to established QRS morphological criteria.^12^ Where a baseline ECG showed ventricular pacing but a LBBB pattern was documented prior to device implantation or at a device check, patients were included as having LBBB.

### Study Endpoints

Follow-up time was calculated from date of first evaluation to date of most recent evaluation, heart transplantation or death from any cause. The primary endpoint was a composite of malignant ventricular arrhythmia (MVA) (defined as sudden cardiac death (SCD), aborted SCD, appropriate implantable cardiac defibrillator (ICD) therapy or sustained ventricular tachycardia (VT)) and end-stage heart failure (ESHF) defined as left ventricular assist device implantation (LVAD), heart transplantation or heart failure-related death.^13^ MVA and ESHF endpoints were analysed separately as secondary endpoints. Patients were censored at the time of their first endpoint event during follow-up or at their last evaluation.

### Statistical Analysis

All data were anonymised and statistical analyses were performed using the Python programming language (Version 3.8, Python Software Foundation, https://www.python.org).^14^ Continuous variables were tested for normality of distribution by visual inspection of histograms and statistical normality tests (Shapiro-Wilk). Normally distributed variables are expressed as mean ± SD and non-normally distributed variables as median [25th, 75th percentiles]. Categorical variables are reported as counts and percentages, as appropriate. The *TableOne* library was used for the construction of summary statistics tables and for all statistical comparisons.^15^ The *Seaborn* and *Matplotlib* libraries were used for data visualization.^16^

The *Lifelines*^17^ library was used for all time-to-event analyses with Kaplan-Meier plots used to display the cumulative probability of the occurrence of endpoints. P-values < 0.05 were considered significant. The log-rank test was used to compare survival. The *zEpid* library was used to calculate incidence rates.^18^

Univariable Cox regression was used to assess the association of baseline variables with endpoints after demonstration that the proportional hazards assumption was supported by the data. Where variables had less than 30% of data missing, a stochastic imputation method was used to allow inclusion in multivariable models. The *Sklearn* library (impute.IterativeImputer) was used to perform a total of 10 imputation rounds before returning the imputations computed during the final round.^19^ A round was a single imputation of each feature with missing values.

Elastic net regularisation was applied with a Cox regression model within the *Scikit-survival*^20^ package to select variables most associated the primary composite endpoint for use in a multivariable Cox model. This method adds a penalty term that uses a combination of two other regression techniques, Least Absolute Shrinkage and Selection Operator (LASSO) regularisation and Ridge Regression. Ridge regression prevents overfitting by minimising the sum of the coefficient squares of the independent variables, while LASSO selects key variables by minimising the absolute value of the coefficients. All variables with non-zero coefficients were reported for the primary composite outcome and those with the highest coefficients were included in a traditional multivariable Cox model for transparency.

To account for cardiac resynchronisation therapy (CRT) occurring during follow-up, a time-varying Cox proportional hazards model was constructed. Follow-up for each patient was split into intervals before and after CRT implantation, with CRT included as a time-varying covariate.

## Results

Six hundred and eighty-six patients with DCM (62.1% male, median [IQR] age 50.2 [37.3, 59.2] years) underwent genetic testing. Five hundred and nine (74.2%) were gene-elusive (no identified LP/P variant in a DCM causing gene) with 491 clinically-affected at baseline. Of 144 patients with LBBB, 133 (92.4%) were gene-elusive.

Forty-three *LMNA* cardiomyopathy (62.8% male, median [IQR} age 49.9 [37.1, 57.9]) and 108 *TTN* cardiomyopathy (70% male, median [IQR] age 48.8 [37.1, 56.4]) patients were identified from the UCL and Barts Heart Centre cardiomyopathy database for comparison.

The total cohort for analysis consisted of 642 patients. Baseline characteristics are shown in Table 1. More gene-elusive patients were probands. More *LMNA* patients had atrial fibrillation or flutter prior to baseline assessment. *LMNA* patients had longer PR intervals at baseline. More gene-elusive patients had LBBB and gene-elusive patients had broader QRS complexes, larger left ventricular internal diameters at end diastole (LVIDd) and lower LVEF.

**Table 1:** Baseline characteristics stratified by genotype. Abbreviations: AF, atrial fibrillation; ECG, electrocardiogram; LA, left atrium; LBBB, left bundle branch block; LVEF, left ventricular ejection fraction; LVIDd, left ventricular internal diameter in diastole; LV LGE, left ventricular late gadolinium enhancement; MVA, malignant ventricular arrhythmia; NYHA, New York Heart Association; RV, right ventricle; TAPSE, tricuspid annular plane systolic excursion; VE, ventricular ectopic.

|  | Missing | Overall | Gene elusive | *LMNA* | *TTN* | P-Value |
| --- | --- | --- | --- | --- | --- | --- |
| n |  | 642 | 491 | 43 | 108 |  |
| Male sex, n (%) | 0 | 407 (63.4) | 310 (63.1) | 27 (62.8) | 70 (64.8) | 0.94 |
| Age at presentation (years), median [Q1,Q3] | 0 | 50.9 [38.7, 60.0] | 51.4 [39.9, 60.5] | 49.9 [37.1, 57.9] | 48.8 [37.1, 56.4] | 0.17 |
| Proband, n (%) | 0 | 546 (85.0) | 452 (92.1) | 23 (53.5) | 71 (65.7) | <0.001 |
| White ethnicity, n (%) | 186 | 292 (64.0) | 211 (62.6) | 29 (74.4) | 52 (65.0) | 0.34 |
| Prior AF or flutter, n (%) | 0 | 94 (14.6) | 61 (12.4) | 15 (34.9) | 18 (16.7) | <0.001 |
| MVA prior to baseline, n (%) | 0 | 25 (3.9) | 20 (4.1) | 3 (7.0) | 2 (1.9) | 0.31 |
| Prior history of heart failure, n (%) | 0 | 176 (27.4) | 131 (26.7) | 7 (16.3) | 38 (35.2) | 0.05 |
| Hypertension, n (%) | 0 | 164 (25.5) | 134 (27.3) | 10 (23.3) | 20 (18.5) | 0.16 |
| Diabetes Mellitus, n (%) | 0 | 91 (14.2) | 70 (14.3) | 3 (7.0) | 18 (16.7) | 0.30 |
| NYHA Class III or IV, n (%) | 0 | 91 (14.2) | 75 (15.3) | 3 (7.0) | 13 (12.0) | 0.26 |
| PR interval (ms), median [Q1,Q3] | 216 | 170 [154,190] | 170 [153,188] | 200 [165,234] | 174 [159,189] | 0.005 |
| QRS duration (ms), median [Q1,Q3] | 137 | 106 [96,133] | 109 [98,141] | 102 [99,120] | 98 [89.8,108.0] | <0.001 |
| Presence of LBBB, n (%) | 85 | 142 (25.5) | 133 (28.1) | 3 (13.0) | 6 (10.0) | 0.004 |
| LV internal diameter (diastole) (mm), median [Q1,Q3] | 78 | 58 [53,63] | 58 [53,64] | 54 [49,60] | 58 [53,61] | 0.004 |
| LV ejection fraction (%), median [Q1,Q3] | 58 | 40 [29,47] | 38 [28,47] | 44 [38,55] | 42 [29,46] | 0.002 |
| LA diameter (mm), mean (SD) | 180 | 40.7 (7.3) | 40.8 (7.4) | 40.0 (6.3) | 40.6 (7.1) | 0.82 |
| RV dilatation, n (%) | 46 | 174 (29.2) | 129 (28.4) | 18 (45.0) | 27 (26.7) | 0.07 |
| TAPSE (mm), median [Q1,Q3] | 269 | 20 [15,23] | 20 [16,23] | 17 [15,22] | 18 [15,24] | 0.67 |
| Hypertrabeculation on echo, n (%) | 115 | 62 (11.8) | 42 (10.1) | 4 (14.3) | 16 (18.8) | 0.07 |
| Number VEs on Holter, median [Q1,Q3] | 268 | 285 [14,2262] | 269 [14,2309] | 897.7 [18, 2280] | 298 [15,2122] | 0.76 |
| Presence LV LGE, n (%) | 116 | 286 (54.4) | 231 (55.3) | 15 (62.5) | 40 (47.6) | 0.31 |

### Clinical Outcomes and Survival Analysis

Six hundred and eleven of 642 patients (491 genotype negative, 43 *LMNA* cardiomyopathy and 108 *TTN* cardiomyopathy) had follow-up data available. Median follow-up time was 48.9 [30.7, 82.5] months.

Supplementary Table 3 shows clinical outcomes occurring at baseline or during follow-up (but not prior to baseline). Compared with *LMNA* cardiomyopathy patients, gene-elusive patients had significantly lower rates of atrial fibrillation, new-onset conduction disease, and ICD implantation during follow-up, with rates similar to those seen in *TTN* cardiomyopathy. Gene-elusive patients had less non-sustained ventricular tachycardia (NSVT) than both *LMNA* and *TTN* cardiomyopathy patients. *LMNA* cardiomyopathy patients had more heart transplantation, heart failure death or death of any cause.

The 49 patients that had MVA at or prior to baseline assessment were excluded from survival analyses for the primary composite and secondary MVA endpoints but were included in the analysis of the secondary ESHF endpoint. Thirty-six of 423 (8.5%) gene-elusive patients had the primary composite endpoint during follow-up versus 14/39 (35.9%) of *LMNA* cardiomyopathy patients and 11/100 (11%) of *TTN* cardiomyopathy patients (log-rank test p- value for *LMNA* vs gene-elusive and for *LMNA* vs *TTN* cardiomyopathy < 0.001 and for *TTN* vs gene-elusive = 0.96; Figure 1).

**Figure 1:**
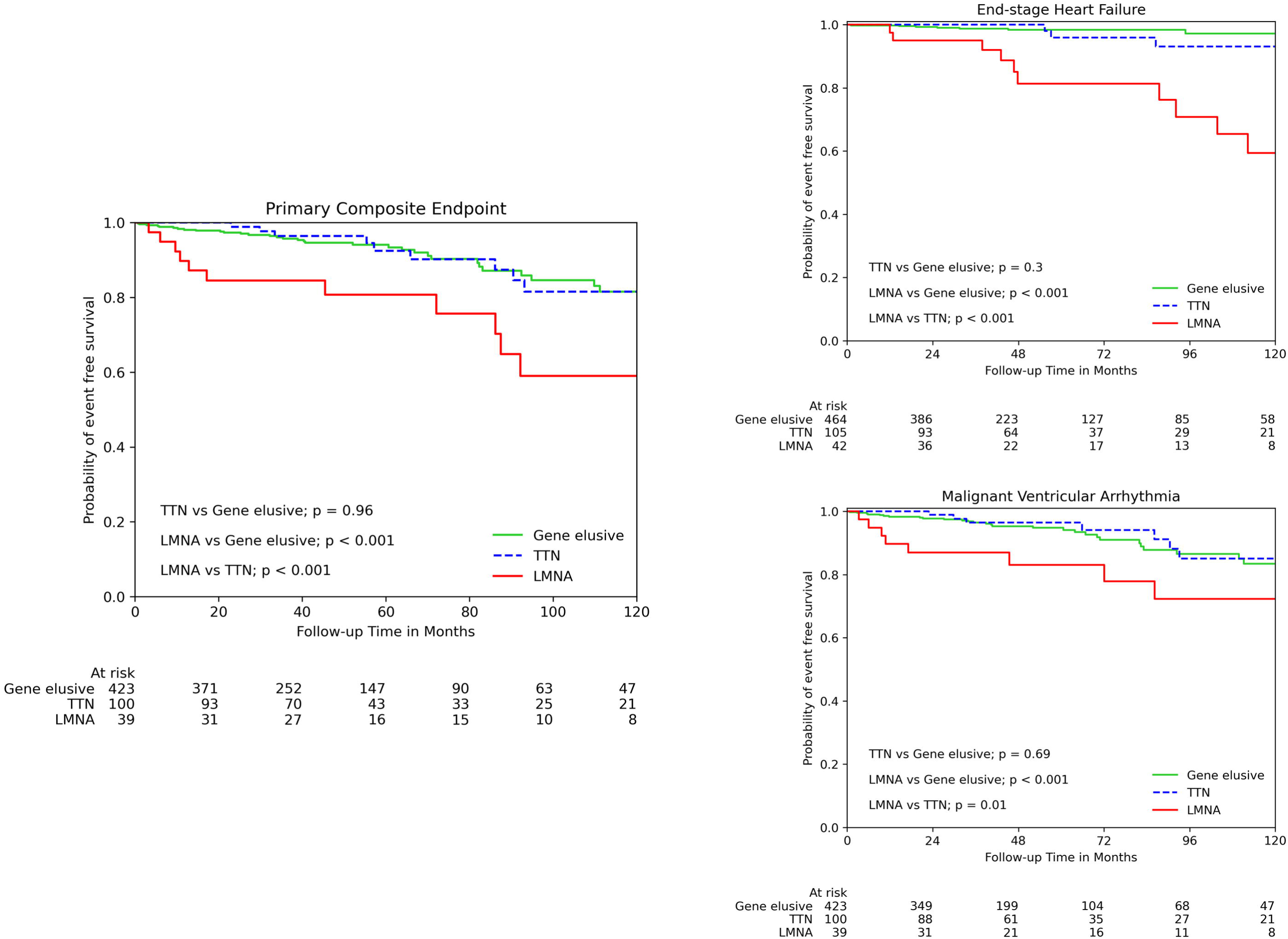
Kaplan Meier survival analysis for a comparison between gene-elusive dilated cardiomyopathy, *TTN* cardiomyopathy and *LMNA* cardiomyopathy patients for a primary composite endpoint of malignant ventricular arrhythmia (MVA) or end-stage heart failure (ESHF) (left) and separate secondary endpoints of ESHF (top right) and MVA (bottom right).

**Figure 2:**
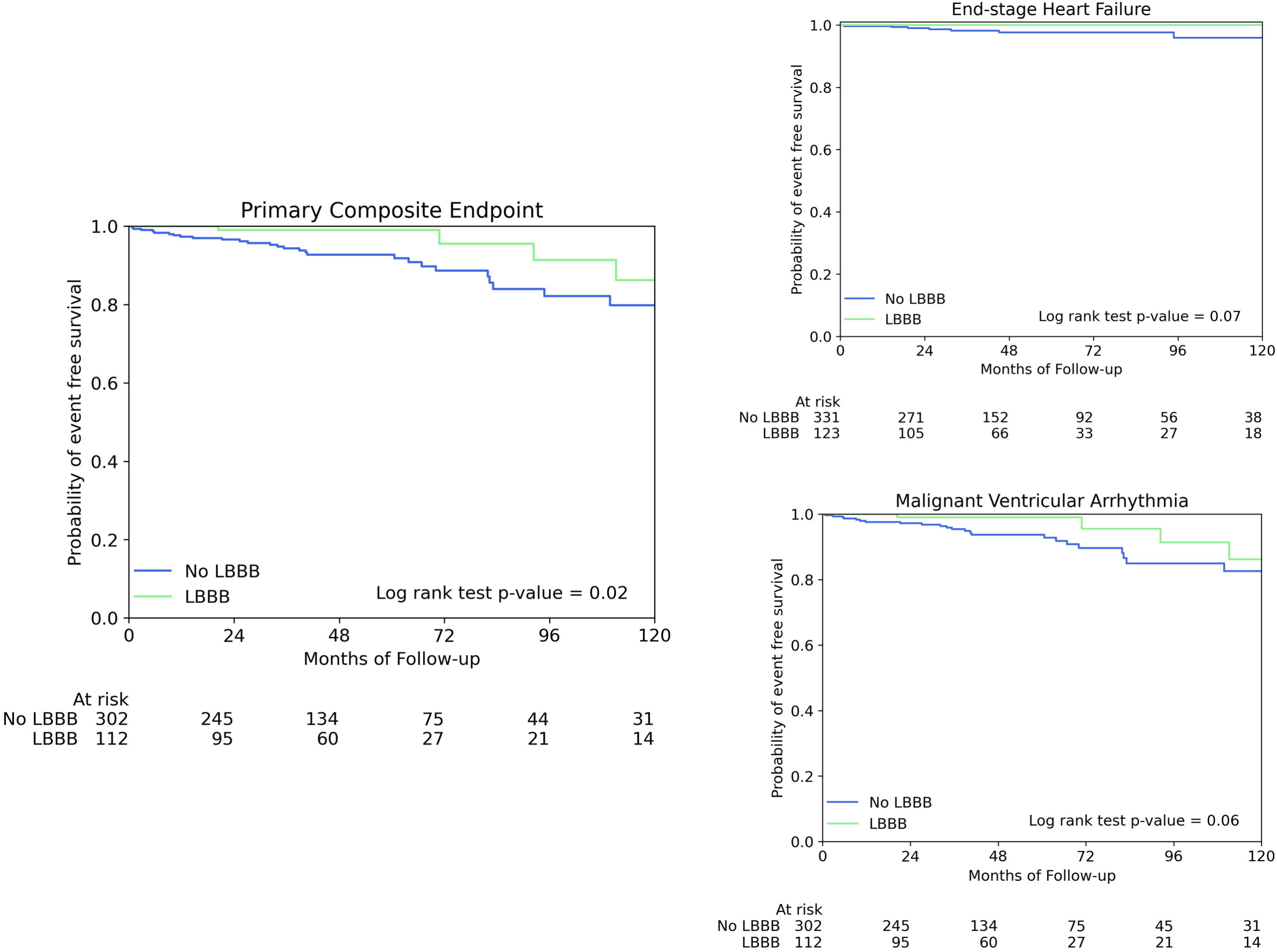
Kaplan Meier Survival analysis for a primary composite endpoint of malignant ventricular arrhythmia (MVA) or end-stage heart failure (ESHF) for gene-elusive dilated cardiomyopathy patients stratified by the presence or absence of LBBB (left) and for the separate secondary endpoints of ESHF (top right) and MVA (bottom right).

Thirty-one of 423 (7.3%) of gene-elusive patients had MVA during follow-up versus 10/39 (25.6%) of LMNA cardiomyopathy and 8/100 (8%) of TTN cardiomyopathy patients (log-rank test p value for LMNA vs gene-elusive < 0.001; LMNA vs TTN = 0.01 and TTN vs gene-elusive 0.69).

Eight from 464 (1.7%) gene-elusive patients had ESHF during follow-up versus 13/42 (31%) *LMNA* cardiomyopathy and 4/101 (4.0%) TTN cardiomyopathy patients (log-rank test p value for *LMNA* vs gene-elusive < 0.001; LMNA vs TTN = <0.001 and TTN vs gene-elusive 0.3).

### Association of baseline variables with outcomes

On univariable analysis of the whole cohort (gene-elusive, *TTN* and *LMNA* cardiomyopathy patients), gene-elusive status showed a trend towards lower risk of the primary composite endpoint (HR 0.6, 95% CI 0.36–1.00; p = 0.05), while *TTN* cardiomyopathy showed no association (HR 0.8, 95% CI 0.3–1.5). In contrast, *LMNA* cardiomyopathy was associated with a significantly higher hazard (HR 3.4, 95% CI 1.8–6.2; p < 0.001).

Within the gene-elusive cohort, univariable Cox regression showed multiple variables associated with the primary composite endpoint including LVIDd, LVEF, LGE on cardiac MRI and ventricular ectopy on ECG (Supplementary Table 4). A family history of cardiomyopathy or of sudden cardiac death did not associate with the primary composite endpoint.

Feature selection for the gene-elusive cohort, using penalised Cox regression within elastic net, showed 5 features with non-zero coefficients associated with the primary composite endpoint (Supplementary Figure 1). A multivariable Cox model using the 4 variables with the highest coefficients showed LVEF, absence of LBBB on ECG, LVIDd and ventricular ectopy on baseline ECG to be independently associated with the primary endpoint (Supplementary Table 4). All variables remained independently associated with the primary endpoint when sex and age at presentation were included in the model.

The addition of the variable ‘CRT at baseline’ to the model attenuated the association between LVIDd and the primary composite endpoint but did not impact the significance of the absence of LBBB at baseline. To examine the effect of CRT during follow-up, a time-varying CRT variable was constructed for inclusion in the model (Supplementary Table 5). LVEF, absence of LBBB on ECG and ventricular ectopy on baseline ECG were independently associated with the primary composite in this model while the time-varying CRT variable was not.

### Left Bundle Branch Block

Four-hundred and seventy-four gene-elusive patients had baseline ECG data available (Table 2). Patients with LBBB were older and more were female. Fewer patients with LBBB had a previous history of atrial fibrillation, but more with LBBB had a previous history of heart failure and CRT at baseline. LVEF was lower in the LBBB group. The median number of ventricular ectopics was higher in those without LBBB.

**Table 2:**
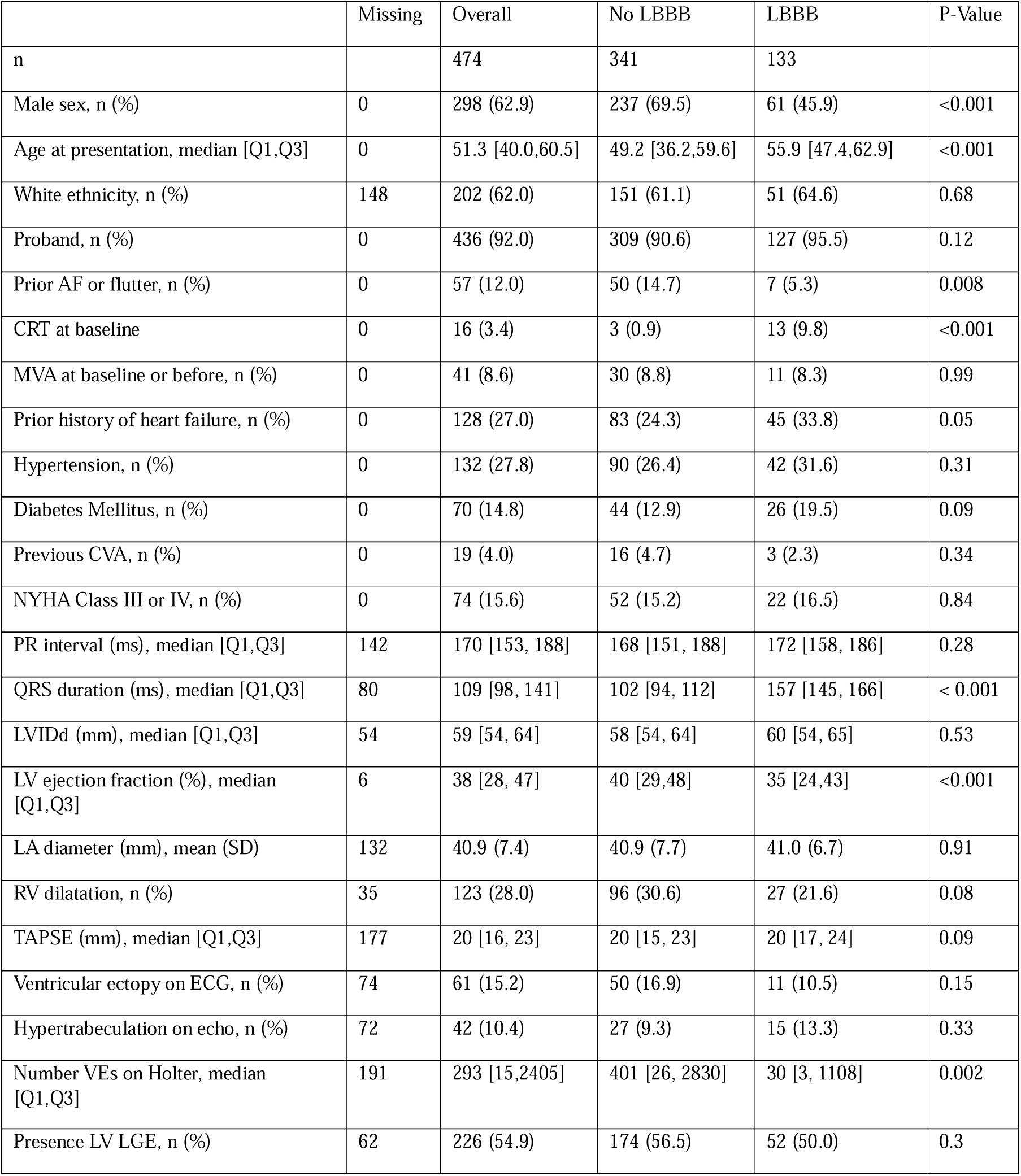
Baseline characteristics of the gene-elusive group stratified by presence or absence of LBBB. Abbreviations: AF, atrial fibrillation; ECG, electrocardiogram; LA, left atrium; LBBB, left bundle branch block; LVEF, left ventricular ejection fraction; LVIDd, left ventricular internal diameter in diastole; LV LGE, left ventricular late gadolinium enhancement; MVA, malignant ventricular arrhythmia; NYHA, New York Heart Association; RV, right ventricle; TAPSE, tricuspid annular plane systolic excursion; VE, ventricular ectopic.

Four-hundred and sixty-four of these gene-elusive patients had follow-up data (Supplementary Table 6). Patients without LBBB had significantly more cerebrovascular events (CVAs) than those with LBBB. Those with LBBB had more ICDs implanted during follow-up but those without LBBB had more appropriate shocks. More of those with LBBB had CRT by the end of follow-up. Sixty-six of 464 patients (14.2%) had ESHF or MVA at baseline or during follow-up (8/464 (1.7%) had an ESHF event and 61/464 (13.1%) had an MVA event).

The 41 patients who had MVA at or prior to baseline assessment were excluded from survival analyses for the primary composite and secondary MVA endpoints but were included in the analysis of the secondary ESHF endpoint. Thirty-five of 414 had the primary composite endpoint during follow-up of 46 [29-75] months; 31/302 (10.3%) without LBBB and 4/112 (3.6%) of those with LBBB (log-rank p value = 0.02; Figure 3). 30 from 414 had the secondary MVA endpoint; 26/302 (8.6%) of those without LBBB and 4/112 of those with LBBB (log-rank test p value = 0.06). Eight from 454 had the secondary ESHF endpoint; 8/331 without LBBB and 0/123 of those with LBBB (log-rank test p value = 0.07).

**Figure 3:**
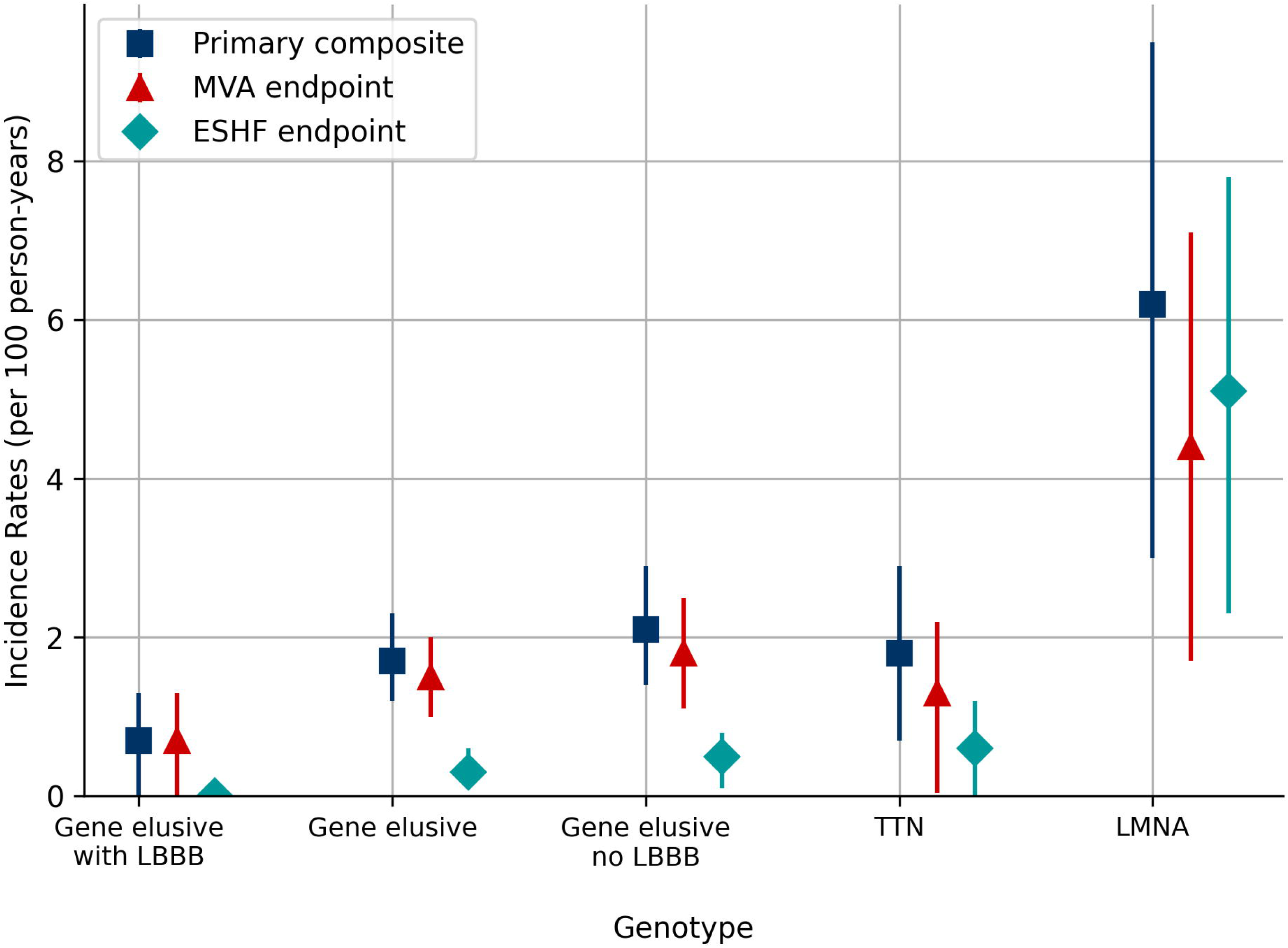
Incidence rates [95% CI] for the primary composite endpoint of malignant ventricular arrhythmia (MVA) or end-stage heart failure (ESHF), as well as for MVA and ESHF separately. Data are stratified by genotype, and within the gene-elusive group, by presence or absence of left bundle branch block (LBBB).

The incidence rate of the primary composite endpoint during follow-up for all gene-elusive patients was 1.7 [1.2 – 2.3] per 100 person years. For genotype negative patients with LBBB the incidence rate was 0.7 [0 – 1.3] per 100 person years and without LBBB it was 2.1 [1.4 – 2.9]. For comparison, the rates in *TTN* and *LMNA* cardiomyopathy patients were 1.8 [0.7 – 2.9] and 6.2 [3.0 – 9.5] per 100 person years, respectively (Figure 3). On univariable Cox analysis within the entire cohort (*LMNA*, *TTN*, gene-elusive), gene-elusive status with LBBB conferred a hazard ratio [95% CI] for the primary composite of 0.3 [0.1-0.8], p-value 0.01).

## Discussion

This study shows that gene-elusive DCM patients have a relatively low risk of adverse events, comparable to that seen in patients with *TTN* cardiomyopathy. Gene-elusive status in combination with baseline LBBB was associated with a favourable prognosis, even after adjustment for CRT implantation modelled as a time-varying covariate.

### Heterogeneity in gene-elusive cardiomyopathy

To our knowledge, there are no studies systematically examining outcomes in gene-elusive DCM, as these patients are typically included only as comparator groups in studies of genotype-positive cohorts. A large Spanish multicentre study, with 691 gene-elusive patients, reported low rates of ESHF and MVA in both gene-elusive and *TTN* cardiomyopathy patients compared to patients with variants in *a priori* designated high-risk genes such as *LMNA*, *FLNC* and *RBM20*.^21^ A single centre Italian study genotyped 487 DCM patients and compared gene-elusive and genotype-positive patients across a range of endpoints.^22^ Strong trends towards worse outcomes were seen in genotype-positive patients for both MVA (p = 0.06) and ESHF (p = 0.06).

In this study, patients with gene-elusive DCM had a better prognosis than those with *LMNA* variants, which are known to be associated with a high risk of adverse events. However, they were still prone to important outcomes including heart failure admissions (15%), appropriate ICD shocks (16%) and myocarditis (4%).

Baseline LVEF, LVIDd, ventricular ectopy on baseline ECG and the absence of LBBB were independently associated with the primary composite endpoint. The association between LV size and systolic dysfunction and adverse outcomes is consistent with observations from ungenotyped, idiopathic DCM populations^23^ however confirmation of these relationships in a gene-elusive cohort is important. Ventricular ectopy has been associated with outcomes in DCM, most notably in the recent *DSP* risk calculator, which incorporates ectopy burden on ambulatory monitoring.^24^ Its identification here highlights the importance of ventricular ectopy in gene-elusive DCM too. The most novel finding is that LBBB on baseline ECG is associated with a better prognosis.

Beyond these markers of risk, clinicians should remain vigilant for additional disease features that may warrant more intensive follow-up or referral to an inherited cardiac disease service for comprehensive risk stratification or expanded genetic testing. Such features may include high-risk patterns of fibrosis on CMR, early-onset conduction disease or atrial arrhythmias, episodes of myocardial inflammation or a malignant family history. Family history was not clearly linked to adverse outcomes in this study, but given the limitations of reporting, and the evolving understanding of genetics, some patients currently classified as gene-elusive will likely receive a genetic diagnosis in the future.

### Left bundle branch block cardiomyopathy as a distinct low-risk phenotype

The proportion of patients with LBBB in our study was two to three-fold higher in the gene-elusive group than either the *LMNA* or *TTN* cardiomyopathy groups. A similar finding was seen in a cohort of Spanish DCM patients and the absence of LBBB was associated with an increased probability of returning a LP/P variant on genetic testing.^25^ The overall yield of genetic testing in our cohort was 25.8%, but only 7.6% in those with LBBB. The low genetic yield in DCM patients with LBBB supports the hypothesis that LBBB cardiomyopathy represents a distinct aetiology of LVSD.

LBBB affects the normal sequence of left ventricular mechanical events in the cardiac cycle to include prolongation of diastolic filling time, abnormal interventricular septal motion and a reduced contribution of the septum to LV ejection fraction.^26^ Patients with intermittent LBBB demonstrate LVSD with the onset of LBBB and recovery of LV systolic function coinciding with the return of normal conduction.^27^ It is, therefore, a reasonable hypothesis that long-standing LBBB could cause a DCM phenotype with a lower risk profile than a DCM caused by a primary myocardial abnormality.

Previous data on the prognostic significance of LBBB in DCM are inconsistent. A cohort of DCM patients were stratified for presence or absence of LBBB at baseline and analysed for an endpoint of all-cause death or heart transplantation.^28^ In contrast to our study, this work identified a poorer prognosis in those with baseline LBBB and in those developing new-onset LBBB during follow-up. The absence of genetic data for this cohort precludes further comparison with our study however the presence of undetected variants in genes associated with a high-risk of adverse events in the LBBB arm of the study may offer an explanation. A meta-analysis of predictors of sustained ventricular arrhythmias in non-ischaemic DCM included seven studies with 1,523 patients and reported a hazard ratio of 1.05 (95% CI: 0.53–2.09) for the presence of LBBB.^29^

The use of CRT has the potential to confound the results of the study of clinical outcomes in patients with LBBB. However, incorporating CRT as a time-varying covariate in multivariable Cox models did not alter the finding that absence of LBBB was associated with poorer prognosis among gene-elusive DCM patients. Even if the lower risk observed in patients with LBBB is partly attributable to the prognostic benefit of CRT, eligibility for CRT is itself a direct consequence of underlying LBBB. This supports the concept of LBBB cardiomyopathy as an aetiologically distinct and lower-risk DCM phenotype.

### Clinical implications of lower-risk phenotypes in gene-elusive DCM

Identification of lower-risk cardiomyopathy patients is increasingly important in the context of over-burdened healthcare systems. Among gene-elusive DCM patients, those with LBBB experienced a particularly low rate of adverse events—less than half that of the gene-elusive cohort overall and approximately three-fold lower than patients without LBBB—despite having lower baseline LVEF. Patients with LBBB also demonstrated a lower burden of ventricular ectopy at baseline, consistently lower rates of atrial arrhythmia and a significantly reduced incidence of stroke. These data suggest that reduced frequency of follow-up, or discharge from specialist care, may be appropriate for stable patients, particularly in the absence of baseline variables associated with the primary composite endpoint.

DCM research to date has largely focused on defining genotype–phenotype relationships, with efforts now pivoting to the development of novel therapies targeting specific genetic mechanisms. While these advances are important, they overlook the significant proportion of patients in whom no pathogenic variant has been detected. Future work should therefore prioritise improved characterisation of the gene-elusive population, to ensure these patients are not excluded from the benefits of precision medicine.

## Limitations

The use of RedCap aimed to mitigate against unstandardised data collection that can confound retrospective studies.^11^ Our centre is a specialist cardiomyopathy unit and thus the study population may be subject to potential referral bias. Although extensive efforts were made to adjust for the influence of CRT implantation on outcomes in gene-elusive patients with LBBB, residual confounding or other unrecognised methodological limitations may have influenced the observed results. Consequently, these findings may not be generalisable to other patient populations.

## Conclusion

Gene-elusive status confers a risk of adverse events comparable to that of *TTN* cardiomyopathy. Baseline factors independently associated with adverse events for gene-elusive patients are lower LVEF, increased left ventricular diameter, ventricular ectopy on ECG and absence of LBBB. The presence of LBBB at baseline identifies a particularly low-risk group that likely represents a distinct aetiology of LVSD, independent of primary myocardial disease.

## Data Availability Statement

Patient level data will not be made available as consent was not sought for public dissemination and due to concerns that information could be used to identify individuals.

