## Supplementary Material for "Characteristics and Outcomes of Gene-Elusive Dilated Cardiomyopathy"

**Supplementary Material: Characteristics and Outcomes of Gene-Elusive Dilated Cardiomyopathy**

*ABCC9, ACTA1, ACTC1, ACTN2, ALMS1, ANKRD1, BAG3, BRAF, CAV3, CRYAB, CSRP3, CTF1, DES, DMD, DNAJC19, DOLK, DSC2, DSG2, DSP, EMD, EYA4, FHL2, FHOD3, FKRP, FKTN, FLNC, FOXD4, GAA, GATA4, GATA6,GATAD1, GLA, GLB1, HFE, JUP, KCNJ2, KCNJ8, LAMA2, LAMA4, LAMP2, LDB3, LMNA, MURC, MYBPC3, MYH6, MYH7, MYL2, MYL3, MYOT, MYPN, NEBL, NEXN, NKX2-5, PDLIM3, PKP2, PLN, PRDM16, PSEN1, PSEN2, PTPN11, RAF1, RBM20, RYR2,SCN5A, SGCA, SGCB, SGCD, SLC22A5, TAZ, TBX20, TCAP, TMEM43, TMPO, TNNC1, TNNI3, TNNT2, TPM1, TTN, TTR, TXNRD2, VCL.*

Supplementary Table 1: Genes included in the 81-gene dilated cardiomyopathy panel. All gene-elusive patients underwent the 81-gene panel as a minimum.

| Gene | DNA | Protein | ACMG criteria | Population frequency |
| --- | --- | --- | --- | --- |
| LMNA | c.448A>G | p.Thr150Ala | LP: PM1, PP2, PM5, PM2_supporting | 0.00000657 |
| LMNA | c.1129C>T | p.Arg377Cys | P: PM1, PP2, PM2, PM5, PS4, PP1_supporting | 0.00000657 |
| LMNA | c.1130G>A | p.Arg377His | P: PS3, PM1, PP2, PM2, PM5, PP3, PS4, PP1_supporting | Not found |
| LMNA | c.1146C>T | p.Gly382= | LP: PS3, PS4, PM2, PP1_supporting | Not found |
| LMNA | c.1150del | p.Glu384Argfs*96 | P: PVS1, PM2_supporting | Not found |
| LMNA | c.1221_1224delCCAG | p.Gln408fs*71 | P: PVS1, PM2_supporting | Not found |
| LMNA | c.1357C>T | p.Arg453Trp | P: PS3, PS2, PM1, PP2, PM5, PP3, PM2_supporting | Not found |
| LMNA | c.1401G>A | p.Trp467* | P: PVS1, PM2_supporting | Not found |
| LMNA | c.1434dupG | p.Leu479Alafs*73 | P: PVS1, PM2_supporting | Not found |
| LMNA | c.1443C>A | p.Trp481* | P: PVS1, PM2_supporting | Not found |
| LMNA | c.1489-1G>A | - | P: PVS1, PM2_supporting | Not found |
| LMNA | c.1567G>A | p.Gly523Arg | LP: PM1, PP2, PM5, PP3, PM2_supporting | 0.0000657 |
| LMNA | c.1634G>A | p.Arg545His | LP: PM1, PP2, PM5, PP3, PS4 | 0.000243 |
| LMNA | c.16C>T | p.Gln6* | P: PVS1, PM2_supporting | Not found |
| LMNA | c.1892dupG | p.Ser632Glnfs*72 | LP: PVS1, PM2_supporting | Not found |
| LMNA | c.1960C>T | p.Arg654* | LP: PVS1, PM2_supporting | Not found |
| LMNA | c.224C>T | p.Ser75Phe | LP: PM1, PP2, PP3, PM2_supporting | Not found |
| LMNA | c.266G>T | p.Arg89Leu | LP: PM1, PP2, PM5, PP3, PM2_supporting | Not found |
| LMNA | c.336_340delGTTTA | p.Phe113Glyfs*12 | LP: PVS1, PM2_supporting | Not found |
| LMNA | c.356+1G>C |  | P: PVS1, PM2_supporting | 0.00000657 |
| LMNA | c.459_462delTGAG | p.Ser153Argfs*23 | LP: PVS1, PM2_supporting | Not found |
| LMNA | c.481G>A | p.Glu161Lys | P: PM1, PP2, PS3, PS4, PP3, PP1_supporting, PM2_supporting | Not found |
| LMNA | c.52_53dup | p.Thr19Profs*78 | P: PVS1, PM2_supporting | Not found |
| LMNA | c.568C>T | p.Arg190Trp | P: PS2, PM5, PM1, PP2, PS3, PP3, PP1 strong, PM2_supporting | 0.00000657 |
| LMNA | c.569G>A | p.Arg190Gln | P: PM5, PM1, PP2, PS3, PP3, PP1_supporting, PM2_supporting | Not found |
| LMNA | c.571delG | p.Val191Trpfs*10 | LP: PVS1, PM2_supporting | Not found |
| LMNA | c.646C>T | p.Arg216Cys | P: PM5, PM1, PP2, PS3, PP3, PP1_supporting, PM2_supporting | 0.00000657 |
| LMNA | c.671C>T | p.Thr22Ile | LP: PM1, PP2, PP3, PM2_supporting | Not found |
| LMNA | c.710T>C | p.Phe237Ser | LP: PM1, PP2, PP3, PM2_supporting | Not found |
| LMNA | c.73C>T | p.Arg25Cys | P: PM5, PM1, PP2, PP3, PM2_supporting | Not found |
| LMNA | c.751dupC | p.Gln251Profs*3 | LP: PVS1, PM2_supporting | Not found |
| LMNA | c.825_832delGCAGTCTG | p.Arg275Serfs*2 | LP: PVS1, PM2_supporting | Not found |
| LMNA | c.871G>A | p.Glu291Lys | P: PM5, PM1, PP2, PP3, PM2_supporting | Not found |
| TTN | c.3351dupA | p.Ser1118Ilefs*21 | P: PVS1, PS4, PM2_supporting | Not found |
| TTN | c.4377del | p.Phe1459Leufs*3 | LP: PVS1, PM2_supporting | Not found |
| TTN | c.9049del | p.Arg3017Glufs*9 | LP: PVS1, PM2_supporting | Not found |
| TTN | c.9377del | p.Lys3126Serfs*17 | LP: PVS1, PM2_supporting | Not found |
| TTN | [c.11346_11347dup](https://mutalyzer.nl/normalizer/NM_001267550.2:c.11346_11347dup) | p.Gly3783Glufs*13 | LP: PVS1, PM2_supporting | Not found |
| TTN | c.12870dup | p.Val4291Serfs*12 | P: PVS1, PM2_supporting | Not found |
| TTN | c.13864dup | p.Ile4622Asnfs*12 | LP: PVS1, PM2_supporting | 0.00000657 |
| TTN | c.19968_19969del | p.Cys6657Tyrfs*26 | LP: PVS1, PM2_supporting | Not found |
| TTN | c.39464-12T>G | - | LP: PVS1, PM2_supporting | Not found |
| TTN | c.39527dup | p.Glu11670Argfs*6 | LP: PVS1, PM2_supporting | Not found |
| TTN | c.42205C>T | p.Arg14069* | P: PVS1, PS4, PM2_supporting | Not found |
| TTN | c.42235C>T | p.Arg14079* | P: PVS1, PS4, PM2_supporting | Not found |
| TTN | c.42825T>G | p.Tyr14275* | LP: PVS1, PM2_supporting | Not found |
| TTN | c.44170_44173del | p.Glu14724Lysfs*39 | LP: PVS1, PM2_supporting | Not found |
| TTN | c.44364del | p.Tyr14789Thrfs*15 | LP: PVS1, PM2_supporting | Not found |
| TTN | c.45307C>T | p.Arg15103* | P: PVS1, PS4, PM2_supporting | Not found |
| TTN | c.45349+1G>T | - | LP: PVS1, PM2_supporting | Not found |
| TTN | c.45564_45567dup | p.Val15190Leufs*18 | LP: PVS1, PM2_supporting | Not found |
| TTN | c.46925_46928del | p.Lys15642Metfs*10 | LP: PVS1, PM2_supporting | Not found |
| TTN | c.47494C>T | p.Arg15832* | P: PVS1, PS4, PM2_supporting | 0.00000659 |
| TTN | c.47697C>A | p.Cys15899* | P: PVS1, PM2_supporting | Not found |
| TTN | c.49648+2del | - | P: PVS1, PS4, PS3, PM2_supporting | 0.0000263 |
| TTN | c.50403del | p.Gly16802Glufs*12 | LP: PVS1, PM2_supporting | Not found |
| TTN | c.51436+1G>T | - | P: PVS1, PS4, PM2_supporting | Not found |
| TTN | c.51667C>T | p.Arg17223* | P: PVS1, PS4, PM2_supporting | Not found |
| TTN | c.52035_52036insTT | p.Leu17346Phefs*4 | P: PVS1, PS4, PM2_supporting, PP1_supporting | Not found |
| TTN | c.52307_52310dup | p.Glu17437Aspfs*2 | P: PVS1, PM2_supporting | Not found |
| TTN | c.53093dup | p.Arg17699Serfs*17 | LP: PVS1, PM2_supporting | Not found |
| TTN | c.53287+1G>T | - | P: PVS1, PS4, PM2_supporting | Not found |
| TTN | c.53647_53649delinsA | p.Trp17883Lysfs*6 | LP: PVS1, PM2_supporting | Not found |
| TTN | c.54120del | p.Gly18041Alafs*44 | LP: PVS1, PM2_supporting | Not found |
| TTN | c.54470_54474del | p.Lys18157Serfs*59 | LP: PVS1, PM2_supporting | Not found |
| TTN | c.54636T>G | p.Tyr18212* | LP: PVS1, PM2_supporting | Not found |
| TTN | c.55854C>A A | p.Cys18618* | LP: PVS1, PM2_supporting | Not found |
| TTN | c.58620del | p.Val19541Phefs*22 | P: PVS1, PS4, PM2_supporting | Not found |
| TTN | c.58732+2T>C | - | P: PVS1, PS4, PM2_supporting | 0.0000131 |
| TTN | c.59205del | p.Glu19735Aspfs*24 | LP: PVS1, PM2_supporting | Not found |
| TTN | c.59864dup | p.Asn19955Lysfs*10 | LP: PVS1, PM2_supporting | Not found |
| TTN | c.59926+1G>A | - | P: PVS1, PS4, PM2_supporting | Not found |
| TTN | c.61876C>T | p. Arg20626* | P: PVS1, PS4, PM2_supporting, PP1_supporting | 0.0000592 |
| TTN | c.62201_62202insC | p.Glu20735* | LP: PVS1, PM2 supporting | Not found |
| TTN | c.62722C>T | p.Arg20908* | LP: PVS1, PM2_supporting | Not found |
| TTN | c.63025C>T | p.Arg21009Ter | P: PVS1, PS4, PM2_supporting, PP1_supporting | Not found |
| TTN | c.63607del | p.Ala21203Profs*44 | LP: PVS1, PM2_supporting | Not found |
| TTN | c.67421del | p.Lys22474Serfs*14 | P: PVS1, PS4, PM2_supporting | Not found |
| TTN | c.67495C>T | p.Arg22499* | P: PVS1, PS4, PM2_supporting | 0.0000395 |
| TTN | c.67665_67668del | p.Pro22556Ilefs*18 | LP: PVS1, PM2_supporting | Not found |
| TTN | c.68449C>T | p.Arg22817* | LP: PVS1, PM2_supporting | 0.00000658 |
| TTN | c.70563dup | p.Glu23522Argfs*10 | LP: PVS1, PM2_supporting | Not found |
| TTN | c.71602C>T | p.Arg23868* | P: PVS1, PS4, PM2_supporting | Not found |
| TTN | c.75138_75141del | p.Lys25046Asnfs*8 | LP: PVS1, PM2_supporting | Not found |
| TTN | c.77149_77150del | p.Leu25717Glufs*6 | LP: PVS1, PM2_supporting | Not found |
| TTN | c.78857_78858del | p.Arg26286Thrfs*16 | LP: PVS1, PM2_supporting | Not found |
| TTN | c.78979C>T | p.Arg26327* | P: PVS1, PS4, PM2_supporting, PP1_supporting | 0.0000132 |
| TTN | c.79894del | p.Glu26632Asnfs*12 | LP: PVS1, PM2_supporting | Not found |
| TTN | c.81365T>G | p.Leu27122* | LP: PVS1, PM2_supporting | Not found |
| TTN | c.83126G>A | p.Trp27709* | P: PVS1, PS4, PM2_supporting | Not found |
| TTN | c.83416C>T | p.Arg27806* | P: PVS1, PS4, PM2_supporting, PP1_supporting | 0.00000658 |
| TTN | c.84388del | p.Cys28130Valfs*44 | LP: PVS1, PM2_supporting | Not found |
| TTN | c.85267C>T | p.Arg28423Ter | P: PVS1, PS4, PM2_supporting | Not found |
| TTN | c.85768C>T | p.Arg28590Ter | P: PVS1, PS4, PM2_supporting, PP1_supporting | Not found |
| TTN | c.86116C>T | p.Arg28706Ter | P: PVS1, PS4, PM2_supporting | 0.00000658 |
| TTN | c.86560delG | p.Val28854Cysfs*4 | LP: PVS1, PM2_supporting | Not found |
| TTN | c.91491del | p.Ile30497Metfs*9 | LP: PVS1, PM2_supporting | Not found |
| TTN | c.91990delA | p.Arg30664Aspfs*6 | LP: PVS1, PM2_supporting | Not found |
| TTN | c.92276_92280del | p.Arg30759Lysfs*17 | LP: PVS1, PM2_supporting | Not found |
| TTN | c.93166C>T | p.Arg31056Ter | P: PVS1, PS4, PM2_supporting | 0.00000403 |
| TTN | c.93479G>A | p.Trp31160* | LP: PVS1, PM2_supporting | Not found |
| TTN | c.95008C>T | p.Arg31670* | P: PVS1, PS4, PM2_supporting | Not found |
| TTN | c.95327del | p.Asn31776Metfs*7 | LP: PVS1, PM2_supporting | Not found |
| TTN | c.95415_95416+2del | - | P: PVS1, PP5, PM2_supporting | Not found |
| TTN | c.95723-3_95723-2del | - | LP: PVS1, PM2_supporting | Not found |
| TTN | c.97475_97476del | p.Glu32492Valfs*18 | LP: PVS1, PM2_supporting | Not found |
| TTN | c.97807A>T | p.Lys32603* | LP: PVS1, PM2_supporting | Not found |
| TTN | c.98299del | p.Arg32767Glyfs*26 | P: PVS1, PS4, PM2_supporting | Not found |
| TTN | c.99821del | p.Asn33274Metfs*26 | LP: PVS1, PM2_supporting | Not found |
| TTN | c.100942_100944delinsT | p.Arg33648Phefs*24 | LP: PVS1, PM2_supporting | Not found |
| TTN | c.101094_101097del | p.Ser33699Metfs*9 | P: PVS1, PS4, PM2_supporting | Not found |
| TTN | c.101665_101668del | p.Val33889* | LP: PVS1, PM2_supporting | Not found |
| TTN | c.103357del | p.Val34453Trpfs*4 | LP: PVS1, PM2_supporting | Not found |
| TTN | c.103705A>T | p.Lys34569* | P: PVS1, PS4, PM2_supporting | Not found |

Supplementary Table 2: Likely pathogenic and pathogenic *LMNA* and *TTN* variants comprising the comparator cohorts and their American College of Medical Genetics (ACMG) classification with supporting criteria. *TTN* and *LMNA* variants were annotated according to the NM_001267550.2 and NM_170707.4 transcripts, respectively.

|  | Overall | Gene Elusive | *LMNA* | *TTN* | P-Value |
| --- | --- | --- | --- | --- | --- |
| n | 611 | 464 | 42 | 105 |  |
| AF or Flutter, n (%) | 161 (26.4) | 110 (23.7) | 20 (47.6) | 31 (29.5) | 0.002 |
| Non-sustained VT, n (%) | 215 (35.2) | 149 (32.1) | 20 (47.6) | 46 (43.8) | 0.02 |
| Cerebrovascular Accident, n (%) | 23 (3.8) | 16 (3.4) | 2 (4.8) | 5 (4.8) | 0.93 |
| Myocarditis, n (%) | 19 (3.1) | 18 (3.9) | 1 (2.4) | 0 | 0.09 |
| ICD implantation, n (%) | 216 (35.4) | 162 (34.9) | 22 (52.4) | 32 (30.5) | 0.04 |
| Appropriate shock, n (%) | 37 (16.1) | 26 (15.6) | 8 (25.8) | 3 (9.4) | 0.18 |
| Anti-tachycardia pacing only, n (%) | 11 (4.8) | 7 (4.2) | 3 (9.7) | 1 (3.1) | 0.02 |
| Inappropriate Shock, n (%) | 17 (7.4) | 11 (6.6) | 2 (6.5) | 4 (12.5) | 0.46 |
| Sustained VT or VF, n (%) | 73 (11.9) | 56 (12.1) | 9 (21.4) | 8 (7.6) | 0.07 |
| Conduction disease during follow-up, n (%) | 18 (2.9) | 7 (1.5) | 9 (21.4) | 2 (1.9) | <0.001 |
| Heart Failure Admission, n (%) | 98 (16.0) | 68 (14.7) | 10 (23.8) | 20 (19.0) | 0.20 |
| LV Assist Device Implantation, n (%) | 4 (0.7) | 3 (0.6) | 1 (2.4) | 0 | 0.37 |
| Heart Transplantation, n (%) | 12 (2.0) | 2 (0.4) | 8 (19.0) | 2 (1.9) | <0.001 |
| Heart Failure Death, n (%) | 11 (1.8) | 4 (0.9) | 5 (11.9) | 2 (1.9) | <0.001 |
| Sudden Cardiac Death, n (%) | 11 (1.8) | 7 (1.5) | 1 (2.4) | 3 (2.9) | 0.43 |
| Death, n (%) | 44 (7.2) | 29 (6.2) | 8 (19.0) | 7 (6.7) | 0.02 |

Supplementary Table 3: Clinical outcomes for dilated cardiomyopathy patients stratified by genotype. Abbreviations: AF, atrial fibrillation; ICD, implantable cardiac defibrillator; LV, left ventricular; VF, ventricular fibrillation; VT, ventricular tachycardia.

| Variable | Univariable HR [95% CI] | Univariable P Value | Multivariable HR [95% CI] | Multivariable p value |
| --- | --- | --- | --- | --- |
| Baseline LVIDd (per 1mm increase on TTE) | 1.1 [1.06 – 1.14] | < 0.001 | 1.07 [1.00 - 1.13] | 0.04 |
| Baseline LVEF (per 5% decrement) | 1.4 [1.2 – 1.6] | < 0.001 | 1.4 [1.2 – 1.7] | < 0.001 |
| LGE on first CMR | 5.9 [2.3 – 15.4] | < 0.001 |  |  |
| Ventricular ectopy on ECG | 4.0 [1.8 – 8.7] | < 0.001 | 4.6 [2.0 – 10.3] | < 0.001 |
| NYHA class 3-4 at baseline | 3.0 [1.5 – 6.0] | 0.002 |  |  |
| Absence LBBB on ECG | 3.8 [1.2 – 11.9] | 0.02 | 6.9 [2.0 – 23.5] | 0.002 |
| Right ventricular dilatation on TTE | 2.3 [1.2 – 4.4] | 0.02 |  |  |
| Baseline LAD (per 1mm increase on TTE) | 1.08 [1.01 – 1.14] | 0.02 |  |  |
| Non-white ethnicity | 2.1 [1.1 – 4.1] | 0.03 |  |  |
| TAPSE (per 1mm decrease) | 1.1 [1.01 – 1.23] | 0.03 |  |  |
| Prior myocarditis | 3.1 [1.04 – 9.1] | 0.04 |  |  |

Supplementary Table 4: Univariable and multivariable Cox models for the primary composite endpoint for gene-elusive dilated cardiomyopathy patients. Abbreviations: CMR, cardiac magenetic resonance; ECG, electrocardiogram; HR, hazard ratio; LAD, left atrial diameter; LBBB, left bundle branch block; LGE, late gadolinium enhancement; LVEF, left ventricular ejection fraction; LVIDd, left ventricular internal diameter at end diastole; NYHA, new york heart failure association; TAPSE; tricuspid annular plane systolic excursion; TTE, transthoracic echocardiogram.

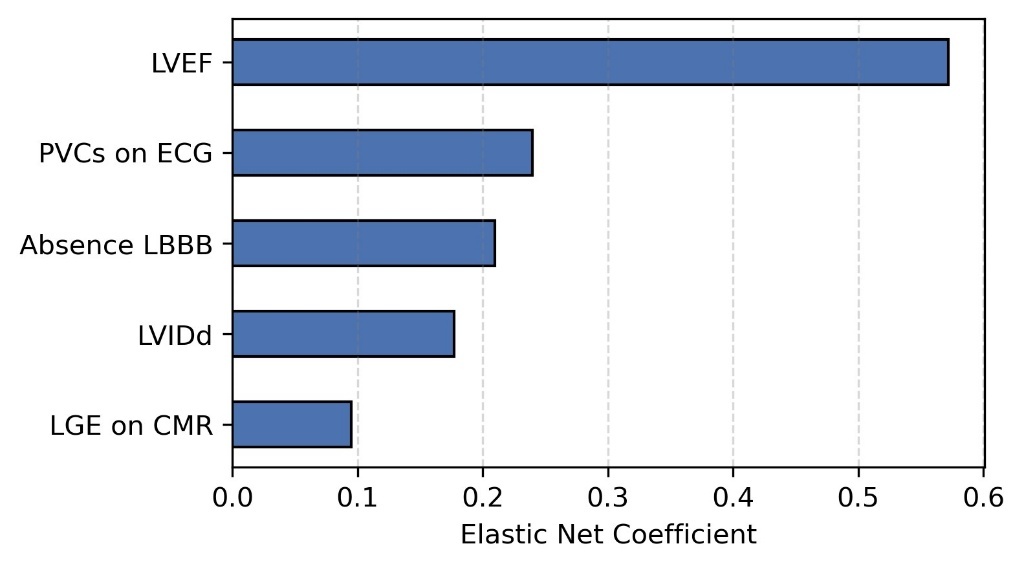

Supplementary Figure 1: Outcome of elastic net feature selection showing variables assessed as non-zero coefficients predictive of the primary composite endpoint for gene-elusive patients using elastic net for feature selection. Abbreviations: CMR, cardiac magnetic resonance scan; ECG, electrocardiogram; LBBB, left bundle branch block; LVEF, left ventricular ejection fraction; LVIDd, left ventricular internal diameter at end diastole; LGE, late gadolinium enhancement; PVC; premature ventricular complex.

| Variable | Multivariable HR [95% CI] | Multivariable p value |
| --- | --- | --- |
| Baseline LVEF (per 5% decrement) | 1.6 [1.4 – 1.8] | < 0.001 |
| Ventricular ectopy on ECG | 3.8 [1.7 – 8.3] | 0.001 |
| Absence LBBB on ECG | 9.5 [2.5 – 36.7] | 0.001 |
| CRT (time-varying) | 2.2 [0.8 – 5.6] | 0.1 |

Supplementary Table 5: A multivariable model for the primary composite endpoint for patients returning no likely pathogenic/pathogenic variants from a DCM gene panel and including a time-varying cardiac resynchronisation variable. Abbreviations: CRT, cardiac resynchronisation therapy; ECG, electrocardiogram; HR, hazard ratio; LBBB, left bundle branch block; LVEF, left ventricular ejection fraction.

|  | Overall | No LBBB | LBBB | P-Value |
| --- | --- | --- | --- | --- |
|  | 464 | 331 | 123 |  |
| AF or Flutter, n (%) | 110 (23.7) | 86 (26.0) | 21 (17.1) | 0.06 |
| Non-sustained VT, n (%) | 149 (32.1) | 109 (32.9) | 36 (29.3) | 0.53 |
| Cerebrovascular Accident, n (%) | 16 (3.4) | 16 (4.8) | 0 | 0.01 |
| ICD implantation, n (%) | 162 (34.9) | 101 (30.5) | 59 (48.0) | 0.001 |
| Appropriate shock, n (%) | 26 (15.6) | 22 (21.6) | 4 (6.6) | 0.02 |
| Anti-tachycardia pacing only, n (%) | 7 (4.2) | 6 (5.9) | 1 (1.6) | 0.26 |
| Inappropriate Shock, n (%) | 11 (6.6) | 10 (9.8) | 1 (1.6) | 0.05 |
| CRT by end follow-up (%) | 78 (16.8) | 20 (6) | 57 (46.3) | <0.001 |
| Sustained VT or VF, n (%) | 56 (12.1) | 45 (13.6) | 10 (8.1) | 0.15 |
| Conduction disease during follow-up, n (%) | 7 (1.5) | 4 (1.2) | 2 (1.6) | 0.66 |
| Heart Failure Admission, n (%) | 68 (14.7) | 53 (16.0) | 14 (11.4) | 0.28 |
| LV Assist Device Implantation, n (%) | 3 (0.6) | 3 (0.9) | 0 | 0.57 |
| Heart Transplantation, n (%) | 2 (0.4) | 2 (0.6) | 0 | 0.99 |
| Heart Failure Death, n (%) | 4 (0.9) | 4 (1.2) | 0 | 0.58 |
| Sudden Cardiac Death, n (%) | 7 (1.5) | 4 (1.2) | 2 (1.6) | 0.67 |
| Death, n (%) | 29 (6.2) | 21 (6.3) | 7 (5.7) | 0.97 |

Supplementary Table 6: Clinical outcomes for the gene-elusive group stratified by presence or absence of left bundle branch block. Abbreviations: AF, atrial fibrillation; CRT, cardiac resynchronisation therapy; ICD, implantable cardioverter defibrillator; LV, left ventricular; LBBB, left bundle branch block; VF, ventricular fibrillation; VT, ventricular tachycardia.
